# Modelling exit strategies for the UK Covid-19 lockdown with revised mortality data

**DOI:** 10.1101/2020.05.05.20091579

**Authors:** Anthony D. Lander, Thejasvi Subramanian

**Affiliations:** Birmingham Women’s and Children’s Hospital, Steelhouse Lane, Birmingham B15 2ES

## Abstract

The number of daily deaths, reported by Public Health England (PHE) during the UK Covid-19 epidemic, initially omitted out-of-hospital deaths in England. The epidemic has been mitigated by social distancing and the lockdown introduced on 17 and 23 March 2020 respectively. We recently reported a stochastic model of a mitigated epidemic which incorporated changes in social interactions and daily movements and whose simulations were consistent with the initial PHE daily mortality data. However, on 29 April, PHE revised their historic data to include out-of-hospital deaths in England. Out-of-hospital deaths occur sooner than in-hospital deaths. Here we show that if 20% of deaths, representing out-of-hospital deaths, are assigned a shorter illness period, then simulated daily mortality matches the revised PHE mortality at least until 4 May. We now predict that if the lockdown is gently relaxed in late May, whilst maintaining social distancing, there would be a modest second-wave which may be acceptable when weighed against the risks of maintaining the lockdown. Our model complements other more sophisticated work currently guiding national policy but which is not presently in the public domain.

---

In an epidemic with fatalities there is a temporal relationship between the size of the infected population, which is unknown, and the subsequent daily deaths which can be ascertained. What may have happened to the infected population can be estimated from what is found in the mortality data. People who die out-of-hospital generally die sooner than those who die in-hospital. Initially, PHE daily mortality data omitted out-of-hospital deaths in England, but included them for other administrations. Since England accounts for 85% of the UK population the initial data disproportionally represented in-hospital deaths which have a longer period from illness to death. On 29 April, PHE revised their historic data and increased total deaths from 21,679 to 26,771^1^. In the revised mortality curve we found changes, which when transposed back 13 days, had a relationship with the introduction of the mitigations, Figure 1.

**Figure 1.**
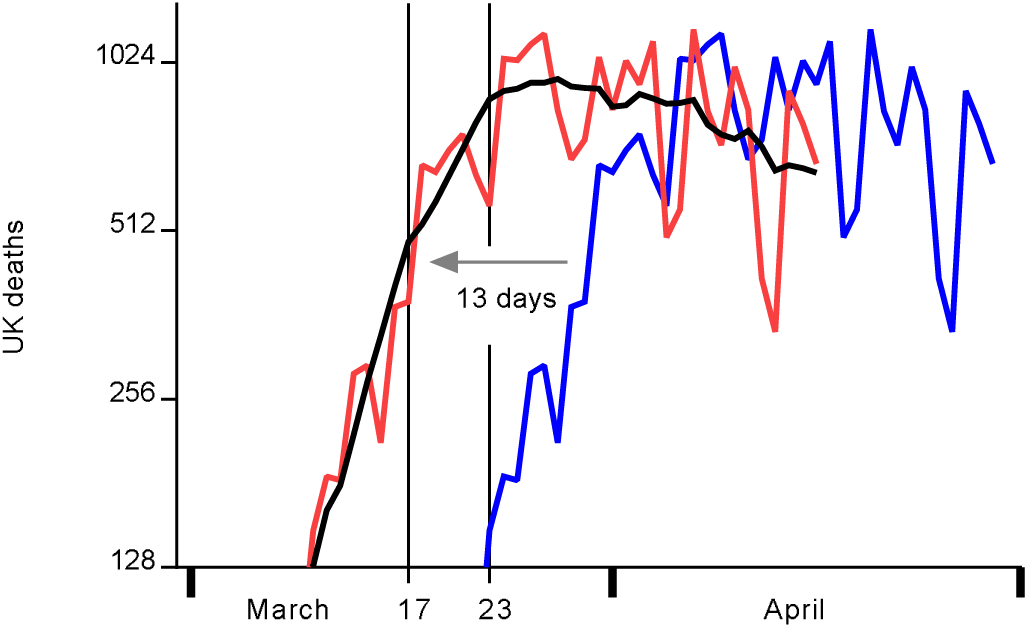
Public Health England’s revised daily deaths (blue) transposed 13 days earlier (red), with a trend line (black) against a log_2_ axis. Social distancing and the lockdown are indicated, 17 and 23 March respectively.

Our model was parameterised as follows; M*_n_* with *n* a surrogate for movements (a larger *n* being more movements), SD*_x_* with x being the percentage of interactions achieving social distancing, and C*_x_* with x being the percentage of symptomatic individuals being confined. Before mitigation, the parameters were (M_18_, SD_10_, C_90_}, on 17 March these changed to {M_14_, SD_50_, C_90_} and on 23 March they changed again to {M_7_, SD_60_, C_100_}. The parameters are detailed here so that the lockdown relaxations, illustrated below, have some context.

In our revised simulations, 20% of deaths were out-of-hospital deaths and were assigned a shorter lognormally distributed interval to death (meanlog 2.639, sdlog 0.2) than the in-hospital deaths (meanlog 2.99, sdlog 0.223). The simulated daily mortality is plotted along with the revised PHE daily mortality in Figure 2.

**Figure 2.**
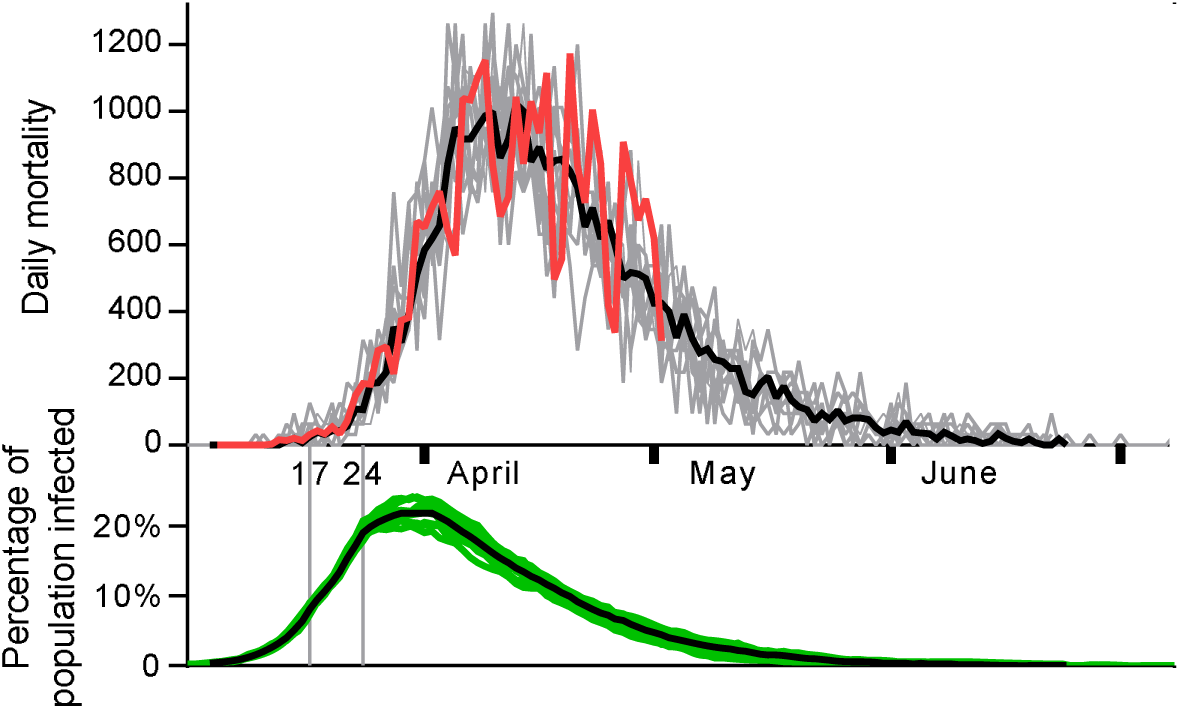
Ten simulations showing the percentage of the population currently infected (green) and daily mortality (grey) with daily means (black). The revised PHE daily mortality is plotted till 4 May (red). Social distancing and lockdown 17 and 24 March are indicated.

The revised PHE figure of 27,510 UK deaths on 1 May coincides with our simulation having recorded 80% of the deaths. This suggests that if the lockdown is maintained, UK deaths will reach a total of 34,000 by early July. After a peak in infections of 23% (20-25%) the epidemic ends, having infected 52% of the population. This leaves a susceptible population at risk of a later epidemic if a sufficient number of contagious individuals were introduced— this could happen at any time. Furthermore, with a population of 66 million this model suggests a UK case fatality rate of 1/1000.

If the lockdown is partially lifted, in May or June, a second-wave will arise, see Figure 3.

**Figure 3.**
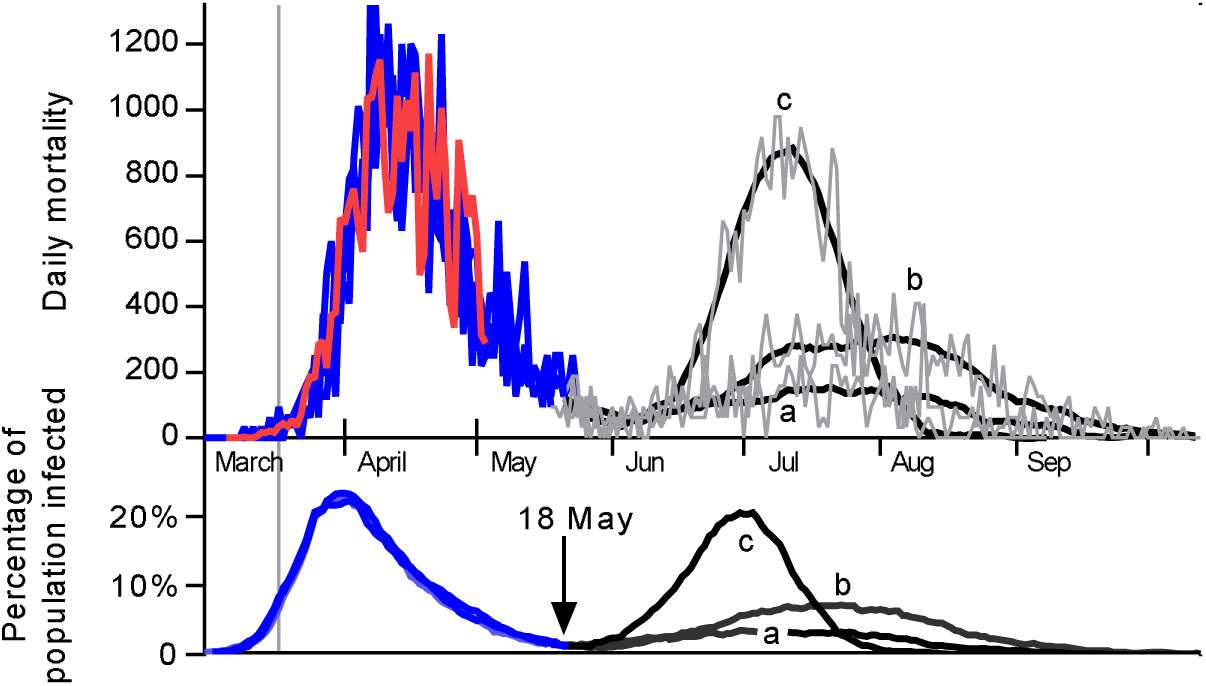
Three simulations showing daily mortality and the percentage of the population infected before relaxation of the lockdown on 18 May (blue). Three simulations of relaxations, introduced on the 18 May, show second-waves in infections and mortalities; (a) {M_14_, SD_50_, C_100_}, (b) {M_20_, SD_50_, C_100_} and (c) {M_18_, SD_10_, C_90_}. Revised PHE UK Covid-19 daily mortality to 4 May (red) is also shown.

The second-wave is more sensitive to changes in social distancing than to changes in the number of movements. The three strategies illustrated in Figure 3 were associated with additional mortalities above the 34,000 of (a) 10,000, (b) 22,000, and (c) 30,000 of which 20% would be out-of-hospital deaths. The three simulations left susceptible populations of (a) 30%, (b) 15%, and (c) 3%. Many of the out-of-hospital deaths are displaced mortalities and this needs to be considered when weighing up the risks and benefits of lockdown release strategies.

## Data Availability

All data is available on-line

## Endnotes

### Author contributions

Anthony Lander developed the stochastic model and wrote the code, Thejasvi Subramanian contributed to analysis and manuscript preparation.

### Additional information

The stochastic model^2^ appears in a preprint awaiting peer review.

### Corresponding author

Correspondence and requests for materials should be addressed to Anthony D. Lander.

### Data availability

Raw data https://www.worldometers.info/coronavirus/

### Code availability

The code used in the stochastic model is available from the corresponding author upon reasonable request.

## Methods

### Stochastic model

A stochastic model was written in Microsoft Excel VBA v7·1 operating in Windows 10. Individuals were assigned an age, sex, risk of mortality if infected, a measure of daily viral exposure, a susceptibility factor, an incubation period, and two contagious periods. One period was for mild illness and the other for serious, critical or fatal illness. Each individual had a 2-dimensional location, a status for symptoms, immunity and a flag for alive/dead. Daily contacts were counted. Individuals had two clocks; one for an assigned contagious period and one for a virtual ‘*twin*’, 80% of whom were assigned a lognormal fatal illness contagious period to represent in-hospital deaths (meanlog 2.99, sdlog 0.223), and 20% a shorter, lognormally distributed period to represent out-of-hospital deaths (meanlog 2.639, sdlog 0.2). Incubation periods were based on Covid-19 data (meanlog 1.621, sdlog 0.418)^3^. Age and sex followed the 2011 UK census. Knowing the age and sex, an individual was designated to die if infected based on a probability taken from early Chinese data, released 11/2/2020 Chinese Centre for Disease Control and Prevention. A random 5% were assigned to be critically ill, and together with the dying individuals, this group had longer contagious (meanlog of 2.99, sdlog of 0.223). A random, normally distributed (mean 1, sd 0.2) susceptibility factor *s* was chosen. Randomly located and randomly moving individuals become infected if they exceed a daily infectious dose based on their cumulative proximity to contagious near neighbours and their susceptibility factor. The exposure, consequent upon an interaction between a susceptible individual *i* and a contagious individual *j*, had a relationship with the inverse square distance separating the individuals *d_i,j_* All separations *d_i,j_* less than 0·2 metres were assumed to be at 0·2 metres. Separations exceeding 5 m were ignored. When social distancing was applied, separations less than 2 m were treated as if at 2 m, this rule was applied for a percentage *x* of interactions designated *SD_x_*. If for any susceptible *i*, on any day, 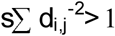 then the individual acquired an infection. One day elapsed between infection and symptoms. A percentage *x*, of symptomatic individuals were confined making no movements and designated *C_x_*, representing isolation. Individuals moved *M_n_* times a day designated M_n_, in a random walk (step-size 10 times the output of an inverse cumulative normal distribution with mean 0 m, sd 10 metres). Individuals returned to their origins at the end of each day. Mobile individuals meeting boundaries were reflected to stay within the boundaries. In these simulations 2000 individuals were placed randomly in a 490 × 490 m square. Only small populations were needed since twins were followed to death (80% having longer and 20% shorter contagious periods. This 80:20 distinguishes the method from that used in doi:10.1101/2020.04.28.20083329. Five contagious individuals were introduced to start an epidemic.

